# Prognostic utility of rhythmic components in 24-hour ambulatory blood pressure monitoring for the risk stratification of chronic kidney disease patients with cardiovascular co-morbidity

**DOI:** 10.1101/2023.05.02.23289413

**Authors:** Nadim El Jamal, Thomas G. Brooks, Jordana Cohen, Raymond R. Townsend, Giselle Rodriguez de Sosa, Vallabh Shah, Robert G. Nelson, Paul E. Drawz, Panduranga Rao, Zeenat Bhat, Alexander Chang, Wei Yang, Garret A. FitzGerald, Carsten Skarke, the CRIC Study Investigators

## Abstract

**Background:** Chronic kidney disease (CKD) represents a significant global burden. Hypertension is a modifiable risk factor for rapid progression of CKD.

**Methods:** We extend the risk stratification by introducing the non-parametric determination of rhythmic components in 24-hour profiles of ambulatory blood pressure monitoring (ABPM) in the African American Study for Kidney Disease and Hypertension (AASK) cohort and the Chronic Renal Insufficiency Cohort (CRIC) using Cox proportional hazards models.

**Results:** We find that rhythmic profiling of BP through JTK_Cycle analysis identifies subgroups of CRIC participants at advanced risk of cardiovascular death. CRIC participants with a history of cardiovascular disease (CVD) and absent cyclic components in their BP profile had at any time a 3.4-times higher risk of cardiovascular death than CVD patients with cyclic components present in their BP profile (HR: 3.38, 95% CI: 1.45-7.88, *p*=0.005). This substantially increased risk was independent of whether ABPM followed a dipping or non-dipping pattern whereby non-dipping or reverse dipping were not significantly associated with cardiovascular death in patients with prior CVD (*p*>0.1). In the AASK cohort, unadjusted models demonstrate a higher risk in reaching end stage renal disease among participants without rhythmic ABPM components (HR:1.80, 95% CI: 1.10-2.96); however, full adjustment abolished this association.

**Conclusions:** This study proposes rhythmic blood pressure components as a novel biomarker to unmask excess risk among CKD patients with prior cardiovascular disease.

## Introduction

Chronic kidney disease (CKD) is a significant global burden, with a prevalence of 9011 cases per 100,000 people. For 2019, this translated into more than 1.4 million deaths globally ^1^. Contributing to that burden is the increasing incidence of major cardiovascular events as kidney function worsens.^2^ Well controlled blood pressure can mitigate these risks, and the International Society of Nephrology blood pressure guidelines suggest the use of ambulatory blood pressure monitoring (ABPM) to complement in-office blood pressure measurements in CKD patients (level 2B recommendation).^3^

A particular advantage of ABPM is the ability to capture blood pressure variability over the course of 24 hours (diurnal rhythms) in a patient’s home environment. Oscillatory signals are abundantly detected in physiologic parameters collected from healthy humans under normal living conditions.^4^ Oscillations are evident in parameters of renal function, including glomerular filtration rate (GFR), renal blood flow, and electrolyte excretion.^5^ Whether deconsolidation of diurnal rhythms contributes to disease expression is of interest in CKD. Loss of renal function leads, for example, to increased sleep fragmentation and conversion of the nocturnal dip in the diurnal blood pressure profile into a non-dipping type which is associated with higher mortality (reviewed by Mohandas et al).^6^ From our analysis of the UK Biobank, the loss of diurnal rhythmicity in wake and sleep periods as assessed by relative amplitude obtained from actigraphy was associated with a higher prevalence of renal failure.^7^

There are many methods to study variability in ABPM recordings. Among these are dispersion methods, methods assessing beat to beat variability and methods quantifying time-specific variations.^8^ Diurnal variability in blood pressure is commonly approached as a binary variable, i.e. presence or absence of the nocturnal dip. The absence of the nocturnal dip, and more so the “inverse dipper” with a rise in blood pressure at night, may be associated with worse renal outcomes in CKD patients.^9^ However, a limitation is that definitions of the nocturnal dip are variable across studies and explore little the characteristics of a time series data set, though comprehensive time-specific assessments are available from adjacent fields. One such method is the JTK_CYCLE algorithm, a non-parametric test to detect the presence of diurnal rhythms primarily in time-integrated gene expression studies.^10^ Here, we adopted the JTK_CYCLE algorithm to parse blood pressure rhythms beyond those detected by the dipping phenotype, and to assess the prognostic utility in ABPM for risk stratification of CKD patients. JTK_CYCLE was designed as a computationally efficient algorithm that detects parameters that cycle across the 24-hour day. This algorithm measures how closely a time series of measurements rises steadily to a peak and descends steadily to a trough. We hypothesized that JTK_CYCLE extracts distinct features in blood pressure time series which are neither picked up by the dipping ratio (which measures the magnitude of decrease in the mean night time blood pressure to the mean day time blood pressure) nor by measures of “beat-to-beat” variability such as average real variability (ARV, which quantifies the dispersion across successive observations). Precedence for the JTK_CYCLE based analysis of time series blood pressure measurements comes from the preclinical domain. Blood pressure rhythmic components were absent in mice lacking the core clock gene BMAL1(brain and muscle ARNT-like protein 1) in their smooth muscles compared to littermate controls, suggesting that peripheral molecular clocks contribute to blood pressure regulation. ^11^

## Methods

### Cohorts

We obtained data from two prospective observational cohort studies of CKD patients; the Chronic Renal Insufficiency Cohort (CRIC) and the African American Study for Hypertension and Kidney Disease (AASK) cohort study. The data were made available upon request from the CRIC publication executive committee and from the NIDDK-CR (a program from the National Institute of Diabetes and Digestive and Kidney Diseases), respectively.

The CRIC study is an ongoing multicenter observational study that enrolled adult patients with various etiologies of chronic kidney disease. Detailed methods have been previously published.^12^ As an ancillary study, participants were randomly chosen from the different clinical sites to undergo ABPM, of whom 1502 provided valid ABPM recordings.^9^ We based our analysis of the CRIC cohort on these participants. Participants who were night-shift workers and who had previous end stage renal disease (ESRD) were excluded and ABPM was set to take measurements every 30 minutes during the day and night. Further details on participant selection and ABPM measurement were previously published.^9^ Outcomes of interest in the CRIC study are a composite cardiovascular outcome of congestive heart failure incidence or events, myocardial infarction, stroke, or peripheral artery disease, cardiovascular death, all cause death, and a composite renal outcome of ESRD or a 50% decline in eGFR. Outcomes were ascertained by asking participants about hospitalizations. Hospital records were then obtained and adjudicated by two clinicians using predefined guidelines.^9^

The rationale and design of the AASK cohort study have been previously published. In brief, The AASK cohort study is a prospective multicenter observational study that is an extension of the AASK trial which tested the effects of a low mean arterial pressure (MAP) goal and the use of antihypertensives on renal function in African Americans with Hypertensive Kidney Disease. In the AASK Cohort study, trial participants who did not reach end-stage renal disease were followed up for five years after trial conclusion with the main outcome of interest being a composite renal outcome defined as a doubling of serum creatinine from baseline, ESRD or Death. A cardiovascular outcome was predefined as a combination of fatal or non fatal cardiovascular events of myocardial infarction, hospitalization for heart failure, or strokes. Outcomes were ascertained from participants at each contact. Hospitalization records were reviewed by a cardiovascular event committee to adjudicate cardiovascular outcomes. Participants had their blood pressure managed by the AASK investigators with a systolic blood pressure goal of less than 130 mmHg and a diastolic blood pressure goal of less than 80 mmHg. Investigators were encouraged to follow a standardized algorithm in their pharmacologic management. 24-hour ABPM with measurements every 30 minutes during both day and night were collected at the post trial baseline and then every other year to total three ABPM measurements over the span of the five years of the study.

### Statistical Analysis

For both studies we considered ABPM recordings with at least 14 readings between 6 am and midnight and at least six readings between midnight and 6am to be valid. ABPM systolic blood pressure values from CRIC and AASK cohorts were binned, for each participant, by the hour of the day and the mean values for each hour were taken. When subjects had more than 24 hours of recording, corresponding hours of all days were binned together. Using Nitecap (nitecap.org),^13^ a web-based tool for the rhythmic analysis of biologic parameters, we ran a JTK_CYCLE analysis on these hourly means. The JTK _CYCLE algorithm is a non-parametric test based on the Kendall tau statistic and operates by performing multiple Jonckheere-Terpstra tests. The Jonckheere-Terpstra test is a non-parametric method similar to the Kruskal-Wallis test but with an assumption of a known ordering of the medians of each group. JTK uses this to compare the actual data’s ordering to the ordering that would have been observed if the data exactly followed a cosine curve. This is then repeated for a library of cosine curves of varying phases followed by a correction for the multiple testing. This results in a P-value that indicates the presence or absence of rhythmic components in the blood pressure curve.^10^ Using this P-value we categorized participants as possessing rhythmic components in blood pressure (JTK P-value ≤0.05) or having no rhythmic components (JTK P-value >0.05).

We calculated dipping ratios (DR) by dividing mean nighttime blood pressure over mean day time blood pressure. Nighttime was defined as a fixed interval between midnight and 6 am. We defined normal nocturnal dipping as a DR less than or equal to 0.9, non-dipping as a DR greater than 0.9 but less than or equal to 1, and reverse dipping as a DR greater than 1. For few analyses, non-dippers and reverse dippers were combined into one group for the interpretability and comparability of results. Assessments of blood pressure control were based on the 2017 ACC/AHA guidelines for blood pressure management.^14^

To assess determinants of either dipping status or the retention of rhythmic components, we fitted multivariable logistic regression models with dipping status or retention of rhythmic components as dependent outcomes. Covariate data were collected at the ABPM visit or the visit closest to that. Because in the AASK cohort study ABPM was repeated every other year, many participants had data from three ABPM sessions. Taking advantage of the multiple measurements for the AASK cohort we ran a mixed effects logistic regression analysis to account for the correlation between measurements from the same participant.

We fitted Cox proportional hazards models to study the utility of the presence of rhythmic components in the risk stratification of chronic kidney disease patients. In this analysis, baseline was determined to be the time of ABPM. Data regarding covariates used in the adjusted models were also collected at either the time of ABPM or the closest visit. For the CRIC cohort, we defined outcomes of interest to be a composite renal outcome (50% decrease in estimated glomerular filtration rate (eGFR) from baseline, or reaching end stage renal disease), death from all causes, death from cardiovascular causes, and a composite cardiovascular outcome (myocardial infarctions, heart failure, strokes, peripheral artery disease). We also fitted Cox proportional hazards models on subgroups stratified by dipping category or the presence of prior CVD. For the AASK cohort, we defined the same outcomes as we did for the CRIC cohort except for the composite cardiovascular outcome which was defined in AASK as cardiovascular death or cardiac revascularization, nonfatal MI, heart failure hospitalization, or stroke.^15^ eGFR was calculated based on the CRIC equation in the CRIC cohort and based on the CKD-EPI 2021 Creatinine equation in the AASK cohort.^16, 17^ Due to the lower event rate and the smaller sample size we did not apply subgroup analyses to the AASK cohort. In survival analyses, data were censored at the end of study follow-up, withdrawal from the study and death if it is not the outcome of interest. Statistical analyses were performed using STATA 17, IBM SPSS version 26, and R 4.2.

## Results

### Cohort characteristics at the time of ABPM

ABPM recordings that passed quality control criteria were available from n=1502 CRIC and n=643 AASK participants. In the AASK cohort, 55.2% (355/643) of the participants completed three valid ABPM sessions that were on average two years apart. A total of 57 CRIC recordings and 16 AASK recordings which did not pass quality control criteria were excluded. The population structure was different for the two cohorts. The CRIC cohort enrolled diverse kidney disease etiologies and aimed for participants with diabetes to account for half of their cohort, while the AASK trial focused only on hypertensive kidney disease patients and excluded those with diabetes in the earlier trial phase. This is reflected by 8.6 mmHg and 10.3 mmHg higher systolic and diastolic blood pressure levels, respectively, in AASK compared to CRIC, and a lower prevalence of diabetes in former. The AASK cohort also had a higher prevalence of CVD, and a mean eGFR which was 10 mL/min lower compared to CRIC (Table 1). Rhythmic components were present in 34% (514/1502) of CRIC and 26% (169/643) of AASK participants (Table S1). Among CRIC and AASK participants categorized as non-dippers, rhythmic components were still detectable in approximately one in seven participants amounting to 16.1% (92/572) and 14.3% (38/266), respectively. We show in Figure S1 blood pressure curves from several CRIC participants satisfying these different patterns. These examples illustrate how time series blood pressure measurements with similar DR and ARV outputs are discriminated by JTK_CYCLE. Two patients with a dipping blood pressure phenotype, DR=0.83 and 0.84 in Figure S1A and Figure S1B, respectively, have a similar ARV of 9.91 and 10.02, respectively, but diverge on the JTK p value of 0.001 compared to 0.36, respectively. Similar juxtaposition is presented for a pair of participants with a non-dipping BP phenotype in Figure S1C and Figure S1D, and for reverse dipping in Figure S1E compared to Figure S1F.

**Table 1.** Characteristics of participants in the two cohorts. Data presented as mean ± S.D. or number (%). The summary statistics are based on data collected at the time of ABPM or the closest visit.

|  | **CRIC(n=1502)** | **AASK (n=643)** |
| --- | --- | --- |
| Clinic Systolic Pressure (mmHg) | 126.23 ± 20.33 | 133.80 ± 20.18 |
| Clinic Diastolic Pressure (mmHg) | 69.24 ± 12.16 | 79.55 ± 11.65 |
| Having Prior CVD | 585 (39%) | 286 (63%) |
| Diabetic | 744 (50%) | 57 (13%) |
| eGFR (mL/min/1.73m<sup>2</sup>) | 46.02 ± 20.31 | 36.06 ± 14.30 |
| Sex (Female) | 661 (44%) | 244(38%) |
| Age (years) | 63.07 ± 10.27 | 60.47 ± 10.13 |
| BMI (Kg/m<sup>2</sup>) | 31.50 ± 6.94 | 31.14 ± 7.01 |
| Race (Black) | 582 (39%) | 643 (100%) |

### Cardiovascular and renal risk factors associated with absence of rhythmic components

Diabetes (*p*=0.007), African American race (*p*=0.03), or a history of cardiovascular disease (*p*=0.001) were associated with absent ABPM rhythmic components in CRIC (Table 2). These covariates did not reach significance in the mixed effects logistic regression applied to the AASK cohort, but BMI greater than 30 (*p*=0.033) was significantly associated here with the absence of rhythmic components in 24-hour blood pressure profiles (Table 3). The association of dipping category and the presence of rhythmic components with different risk factors further indicates these measures capture different populations and set forth the expectation that they might have different associations with health outcomes.

**Table 2.** Multivariate logistic regression for the absence of rhythmic components in blood pressure among CRIC cohort participants. All variables in the table were included as covariates in the model. eGFR: estimated glomerular filtration rate by the CRIC cohort equation. BP: blood pressure. PCR: urine protein to creatinine ratio.

| **Covariate** | **Odds Ratio** | **95% Lower** | **CI 95% CI Upper** | **P-value** |
| --- | --- | --- | --- | --- |
| Age <45 | Reference |  |  |  |
| Age From 45 to <65 | 1.27 | 0.78 | 2.07 | 0.329 |
| Age 65+ | 1.47 | 0.9 | 2.41 | 0.123 |
| BMI <25 | Reference |  |  |  |
| BMI 25 to <30 | 0.87 | 0.61 | 1.26 | 0.465 |
| BMI ≥30 | 1.24 | 0.71 | 1.76 | 0.233 |
| eGFR >60 ml/min/1.73m<sup>2</sup> | Reference |  |  |  |
| eGFR: 30 to <60 ml/min/1.73m<sup>2</sup> | 1.05 | 0.77 | 1.42 | 0.769 |
| eGFR<30 ml/min/1.73m<sup>2</sup> | 1.01 | 0.74 | 1.63 | 0.65 |
| Proteinuria (PCR <150mg/g) | Reference |  |  |  |
| Proteinuria (PCR: 150-500 mg/g) | 1.19 | 0.86 | 1.63 | 0.29 |
| Proteinuria (PCR>500 mg/g) | 1.23 | 0.88 | 1.74 | 0.231 |
| Controlled BP | Reference |  |  |  |
| Un-Controlled BP (Mean 24 Hr BP ≥ 125/75mmHg) | 0.97 | 0.74 | 1.27 | 0.823 |
| Male Sex | Reference |  |  |  |
| Female Sex | 1.002 | 0.78 | 1.28 | 0.988 |
| Non-Diabetic | Reference |  |  |  |
| Diabetic | 1.41 | 1.1 | 1.82 | 0.007 |
| Race: White | Reference |  |  |  |
| Race: Black | 1.5 | 1.15 | 1.96 | 0.03 |
| Race: Other | 1.26 | 0.86 | 1.83 | 0.237 |
| Prior CVD: No | Reference |  |  |  |
| Prior CVD: Yes | 1.58 | 1.22 | 2.05 | 0.001 |

**Table 3.**
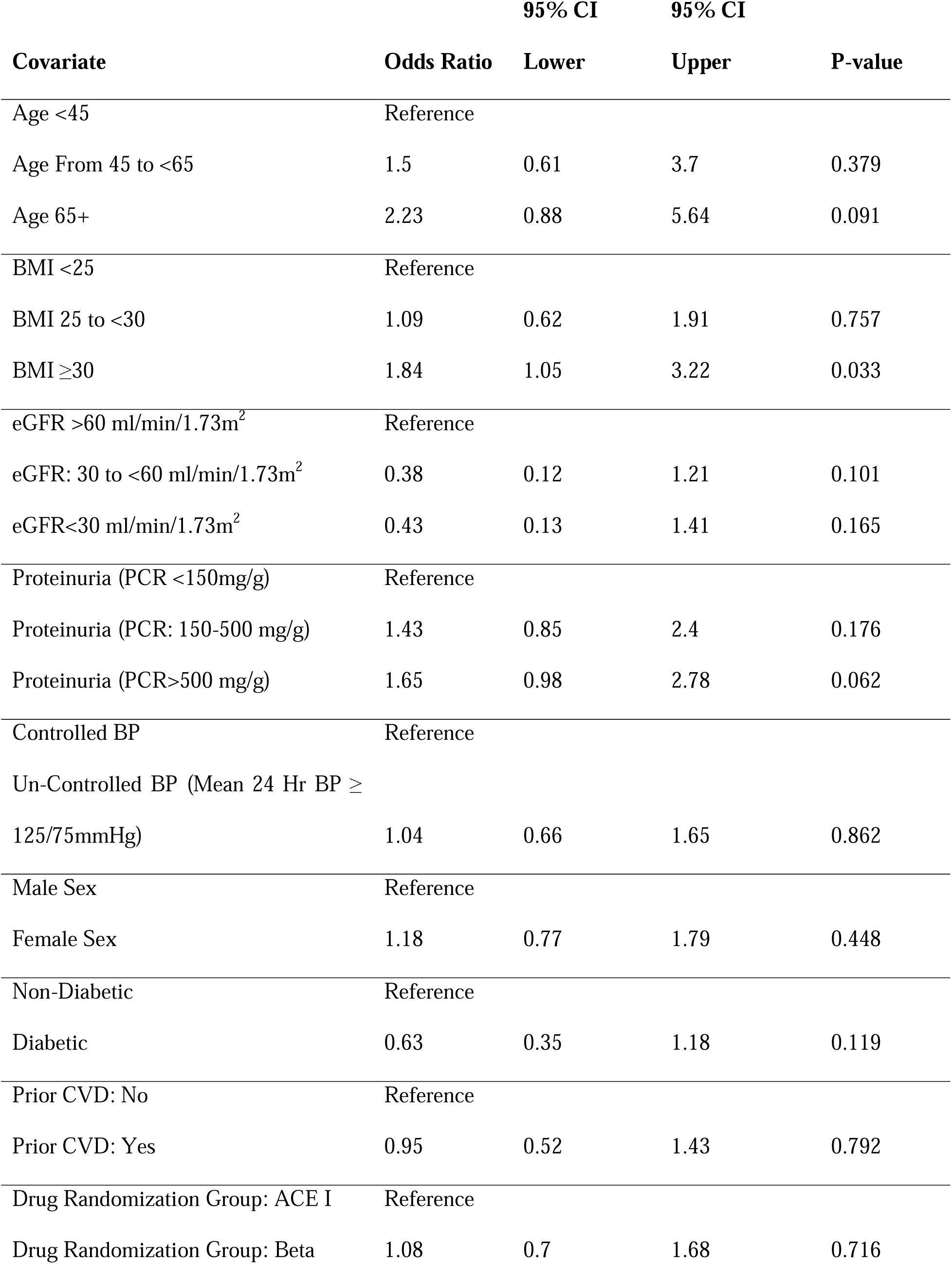
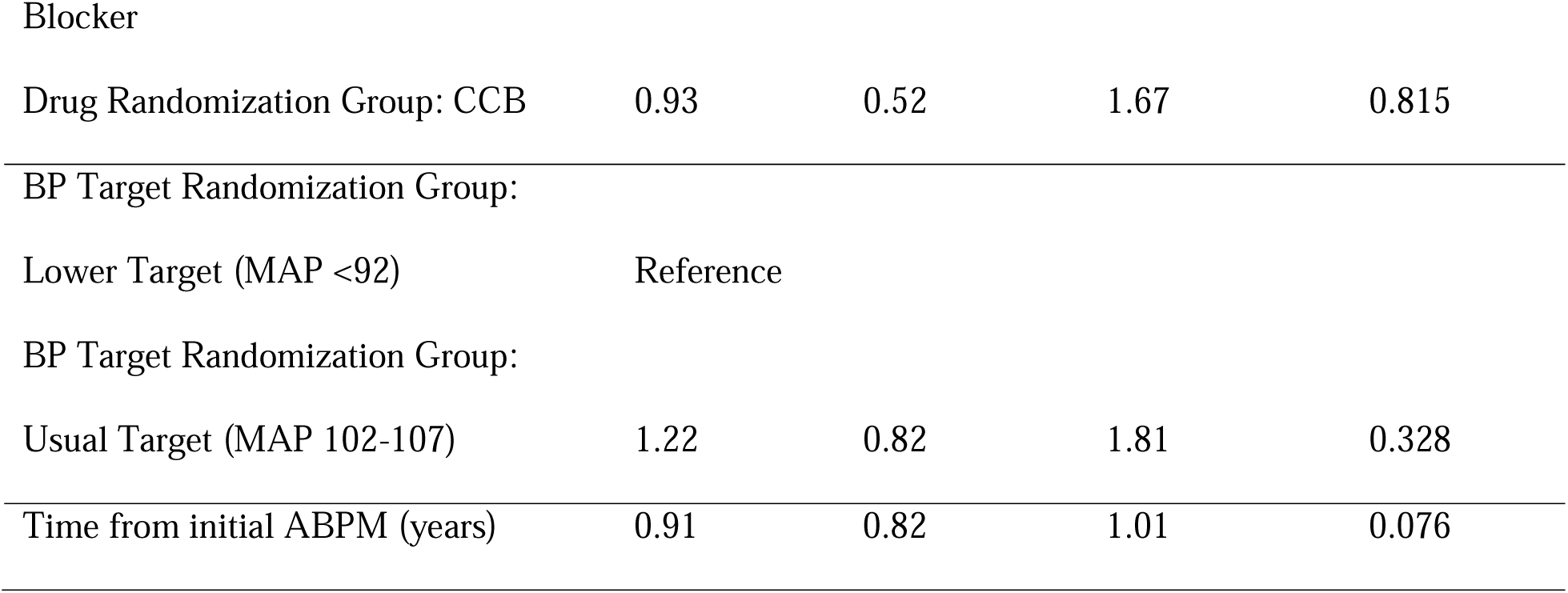
Mixed effects logistic regression for the absence of rhythmic components in blood pressure among AASK cohort participants. All variables in the table were included as covariates in the model. eGFR: estimated glomerular filtration rate by the 2021 CKD-EPI equation. PCR: urine protein to creatinine ratio. BP: blood pressure. ACE I: angiotensin converting enzyme inhibitor. CCB: calcium channel blocker.

### Cardiovascular and renal risk factors associated with non-dipping BP

In the CRIC cohort, variables associated with nocturnal non-dipping included a protein-creatinine ratio (PCR)>500 mg/g (*p*=0.009), uncontrolled BP (*p*<0.001), African American race (*p*=0.001), and a history of cardiovascular disease (*p*<0.001) (Table S2). A trend towards age-dependent emergence of nocturnal non-dipping was noticeable in participants older than 65 years (*p*=0.047). Only uncontrolled BP (*p*=0.006) and taking beta-blockers as anti-hypertensives during the clinical trial phase (*p*=0.016) attained significance in the AASK cohort (Table S3). A history of diabetes (*p*=0.048) trended to be associated.

### Absence of rhythmic BP components as an independent risk factor for cardiovascular death in CKD patients with prior cardiovascular disease

Clinical outcomes were collected over a mean follow-up time of 8.4 (±2.6) years for the CRIC cohort and 4.5 (±1.1) years for the AASK cohort. CRIC participants with a history of CVD and absent cyclic components in their BP profile had at any time a 3.4-times higher risk to reach the outcome of cardiovascular death than participants with cyclic components present in their BP profile (HR: 3.38, 95% CI: 1.45-7.88, *p*=0.005, Figure 1A). This substantially increased risk was not driven by a specific subgroup, i.e. nocturnal dipper, non-dipper or reverse dipper (p>0.1, Table 4). Rhythmic BP components were absent in all non-and reverse-dipping CRIC participants with cardiovascular death (n=24 and n=21, respectively). Among dippers with prior CVD succumbing to cardiovascular death, the majority (14 out of 20) had no cyclic components in their BP profiles, while 6 retained BP cycles (Table S4). We found that the signal was strongest in participants with prior CVD, as the hazard ratios for cardiovascular death were lower in the unadjusted (HR: 2.32, 95% CI: 1.37-3.93, *p*=0.002, Figure S2.A) and adjusted (HR: 1.7, 95% CI: 0.98-2.95, *p*=0.059, Figure S2.B) models, the latter controlling for all covariates including prior cardiovascular disease. Furthermore the interaction term between the absence of rhythmic components and the presence of cardiovascular disease had an adjusted hazard ratio of 4.19 (95% CI: 1.34-14.5, *p=*0.014)

**Figure 1.**
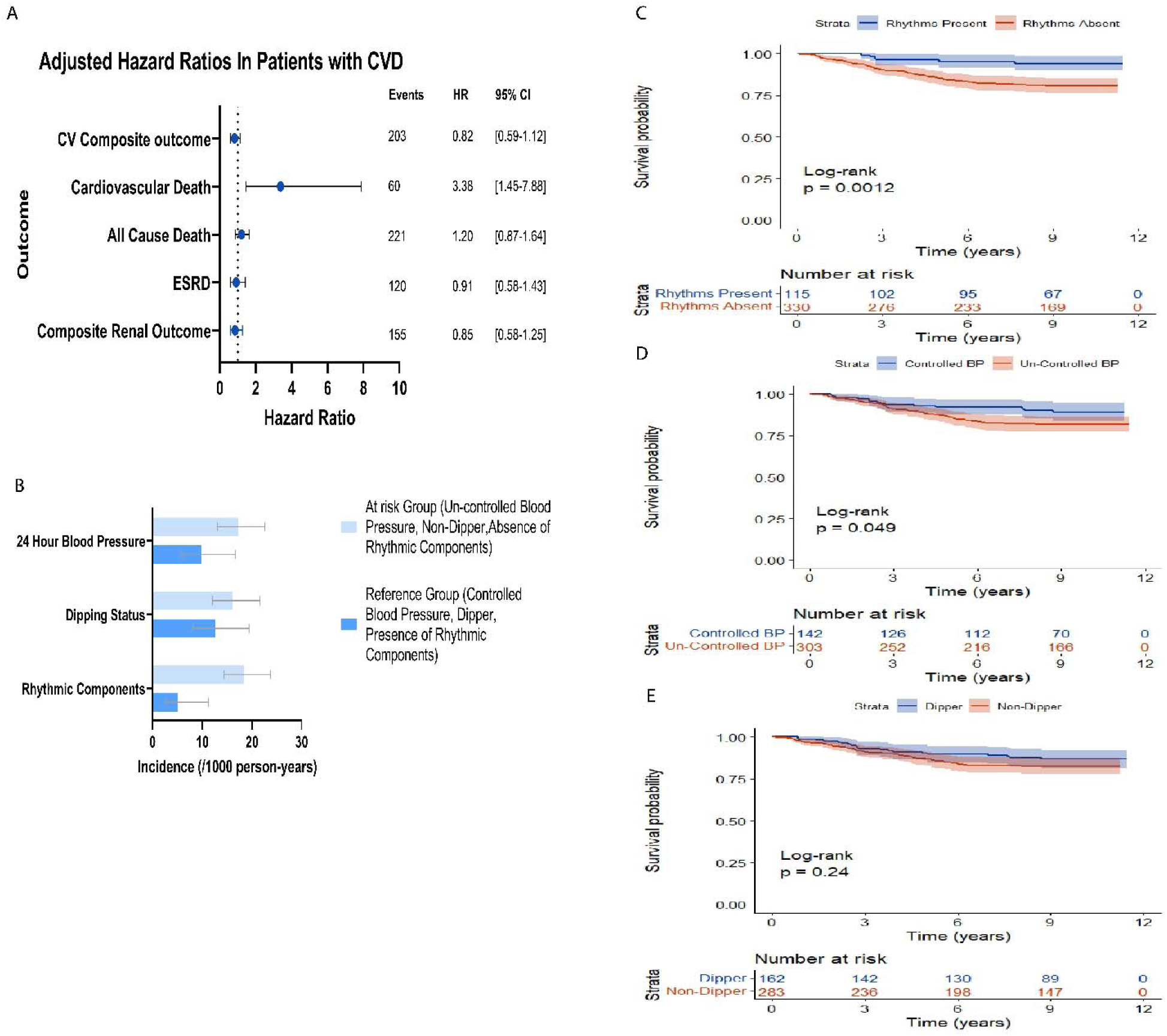
**A:** Hazard ratios for the absence of rhythmic components (JTK p value >0.05) and reaching different outcomes in the CRIC cohort as compared to the retention of rhythmic components (JTK p value ≤0.05) in participant with prior CVD. Adjusted for Age, BMI, Sex, Diabetes, Race, eGFR, Urine Protein to Creatinine Ratio, Clinic SBP, and Clinic DBP. **B:** Incidence of cardiovascular mortality in patients with prior cardiovascular disease stratified by the different ambulatory blood pressure monitoring derived parameters. Kaplan Meier survival estimates for cardiovascular death in CRIC patients with prior CVD stratified by categorizations of the different ambulatory blood pressure parameters. **C.** stratified by the presence of rhythmic components. **D.** stratified by blood pressure control (24 hour blood pressure mean cutoff of 125/75). **E.** Stratified by dipping status (Dipping ratio cutoff of 0.9)

**Table 4.** Hazard ratios for the different dipping categories and reaching different outcomes among CRIC cohort participants with prior CVD Adjusted for Age, BMI, Sex, Diabetes, Race, eGFR, Urine Protein to Creatinine Ratio, Clinic SBP, Clinic DBP. DR: Dipping ratio, CV: Cardiovascular, CVD: Cardiovascular disease, ESRD: End stage renal disease.

| Outcome | Dipping Category | Unadjusted Model |  |  | Adjusted Model |  |  |
| --- | --- | --- | --- | --- | --- | --- | --- |
|  |  | Hazard Ratio | 95% CI | p-value | Hazard Ratio | 95% CI | p-value |
| **Composite Renal Outcome** | Dipper | Reference |  |  |  |  |  |
|  | Non-Dipper | 1.63 | 1.14-2.34 | 0.008 | 1.13 | 0.75-1.70 | 0.545 |
|  | Reverse Dipper | 2.06 | 1.39-3.07 | <0.001 | 1.53 | 0.99-2.38 | 0.058 |
| **ESRD** | Dipper | Reference |  |  |  |  |  |
|  | Non-Dipper | 1.79 | 1.19-2.72 | 0.006 | 0.97 | 0.60-1.57 | 0.893 |
|  | Reverse Dipper | 2.05 | 1.30-3.25 | 0.002 | 1.37 | 0.83-2.28 | 0.217 |
| **All Cause Death** | Dipper | Reference |  |  |  |  |  |
|  | Non-Dipper | 1.01 | 0.75-1.34 | 0.995 | 0.80 | 0.58-1.09 | 0.161 |
|  | Reverse Dipper | 1.48 | 1.09-2.02 | 0.012 | 1.03 | 0.72-1.46 | 0.874 |
| **Cardiovascular Death** | Dipper | Reference |  |  |  |  |  |
|  | Non-Dipper | 1.06 | 0.59-1.92 | 0.843 | 1.07 | 0.57-1.98 | 0.836 |
|  | Reverse Dipper | 1.66 | 0.90-3.07 | 0.104 | 1.29 | 0.64-2.57 | 0.476 |
| **Composite CV outcome** | Dipper | Reference |  |  |  |  |  |
|  | Non-Dipper | 0.93 | 0.69-1.25 | 0.616 | 0.80 | 0.58-1.11 | 0.180 |
|  | Reverse Dipper | 1.18 | 0.85-1.64 | 0.337 | 0.88 | 0.61-1.28 | 0.502 |

To quantify the occurrence of CV death further, we calculated the person-time rate for cardiovascular mortality among CRIC participants with prior CVD (Figure 1B). The absence of rhythmic components had a statistically significant increase in the incidence rate compared to their presence (18.46 [14.30-23.82] per 1000 person years compared to 5.04 [2.65-11.22] per 1000 person years respectively, *p*<0.001). This was neither seen in 24 hour blood pressure control (17.20 [13.07-22.63] per 1000 person years in uncontrolled BP, 9.85 [5.83-22.63] per 1000 person years in controlled BP, *p*=057) nor in dipping category (16.11 [12.03-21.58] per 1000 person years in non-dippers, 12.55 [8.10-19.45] per 1000 person years in dippers, *p*=0.356). In the time-to-event analysis, survival probability was significantly lower in participants without rhythmic BP components compared to those who had them (log-rank *p*=0.0012). Survival probability trended to be lower in participants with uncontrolled BP compared to controlled BP (log-rank *p*=0.049), while no difference was detected for non-dippers compared to dippers (log-rank *p*=0.24) as shown in Figure 1C-E.

We turned to the 83 cardiovascular deaths adjudicated for the adjusted model (compared to 60 in the adjusted model restricted to CVD, Figure S2.B) to explore further whether absence of rhythmic components confers an increased risk for cardiovascular death. Strikingly, we found that reverse dippers without cyclic components trended to be at an 8-fold higher risk of dying from cardiovascular causes compared to reverse dippers with retained rhythms (HR: 8.0, 95% CI: 0.99-63.8, *p*=0.05, Figure S3). While the trend is large, the low number of events and the wide confidence interval slightly intersecting the line of no-effect invite caution in interpreting this result. All cause death emerged as a significant signal (HR: 2.74, 95% CI: 1.31-5.75, *p*=0.007). Absence of rhythmic components had little effect on cardiovascular or all-cause mortality among dippers (HR:1.3, 95% CI: 0.6-2.83, *p*=0.506, and HR: 0.77, 95% CI: 0.59-1.11, *p*=0.158, respectively) and non-dippers (HR:1.51, 95% CI: 0.45-5.04, *p*=0.504, and HR: 1.25, 95% CI: 0.75-2.09, *p*=0.389, respectively). As was previously published,^9^ participants with a non-dipping pattern (particularly reverse dipping pattern) were at a higher risk for worsening kidney function (Table S5).

For the AASK cohort, we detected in the unadjusted Cox proportional hazard model a significant association between the absence of rhythmic BP components and end stage renal disease (HR:1.80, 95% CI: 1.10-2.96, *p*=0.020) as well as the composite renal outcome (HR: 1.48, 95% CI: 1.01-2.16, *p*=0.042). However, these associations lost statistical significance when fully adjusting the model (HR:1.67, 95% CI: 0.73-3.84, *p*=0.227 and HR:1.2, 95% CI: 0.68-2.1, *p*=0.528, respectively, Figure 2). There were no statistically significant associations between dipping status and all outcomes in the AASK cohort except for reverse dipping and the composite cardiovascular outcome in the unadjusted model (HR: 1.79, 95%CI: 1.02-3.14, P-value: 0.043) (Table S6). Due to the smaller sample size and the low number of events, we did not perform subgroup analyses using the AASK cohort data.

**Figure 2.**
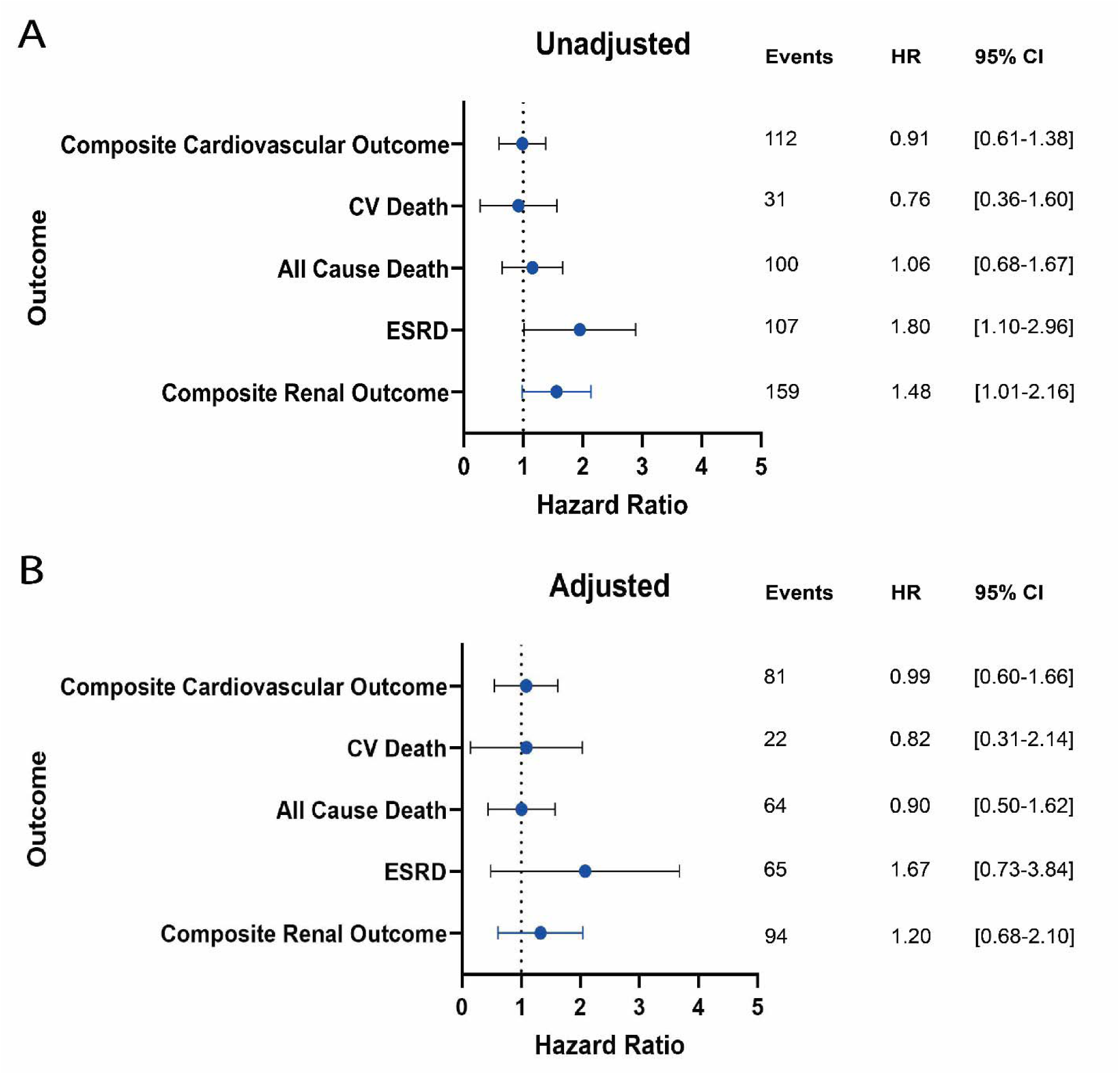
Hazard ratios for the absence of rhythmic components (JTK p value >0.05) and reaching different outcomes in the AASK cohort as compared to the retention of rhythmic components (JTK p value ≤0.05). **A.** Unadjusted model **B.** Adjusted for Age, BMI, Sex, Diabetes, eGFR, Urine Protein to Creatinine Ratio, Clinic SBP, Clinic DBP, Prior CVD, Drug and blood pressure target groups randomized to in the prior trial.

To compare rhythmic components with non-diurnal methods of variability we fitted cox proportional hazard models on tertials of average real variability (ARV), a measure of sequential beat-to-beat variability. We divided the population over three tertials: ARV ≤9(n=579), 9<ARV ≤11 (n=344), or ARV >11 (n=579). We found a significant association between higher ARV tertials and all outcomes except cardiovascular death in the unadjusted model. These associations did not retain statistical significance after full adjustment (Table S7).

## Discussion

The main finding of the present study is that nonparametric characterization of time-dependent effects in ambulatory blood pressure measurements (ABPM) identified patients with chronic kidney disease at increased risk of mortality irrespective of their blood pressure (BP) dipping behavior defined as nocturnal dipper, non-dipper or reverse dipper. This was evident among CRIC participants with a history of CVD where the participants without rhythmic BP components had at any time a 3.4-times higher risk to experience cardiovascular death than the participants with existing rhythmic BP components. We also found that risk factors known to increase cardiovascular and renal adverse events were associated, as expected, with nocturnal non-dipping behavior.^9, 18, 19^ Predictors of rhythmic components and dipping patterns are different and overlap only in demography and prior comorbidities, further suggesting that these metrics capture different population structures. While a study on a smaller sample of CKD patients (470) showed significantly higher risk for all cause death, cardiovascular events, and more rapid renal function decline with a high ARV,^20^ our adjusted model does not show similar results.

The major benefit of quantifying rhythmic components per JTK_CYCLE from BP time series data is its independence from collecting accurate sleep onset and offset data. This non-parametric test has been widely adopted in the circadian clock field to identify oscillating molecular biomarkers,^10^ thus underscoring the reliability to discover cyclical ordering in timed BP readings. Of the blood pressure determinants listed in Gumz M et al.,^21^ each single one has a diurnal preference tied to mechanisms under circadian control. Signals from the suprachiasmatic nuclei (SCN), entrained to the 24-hour environmental cycle, synchronize tissue-specific circadian patterns of clock-controlled gene (CCG) expression through transcriptional-translational loops in the endothelium, vascular smooth muscle cells and the heart. Oscillating Dbp (Albumin D-site-Binding Protein) and Glut4 (glucose transporter 4) gene expression levels drive diurnal fluctuations in vasodilation and heart rate both modulating the diurnal blood pressure phenotype (reviewed in Hastings MH et al.).^22^ Other sources of diurnal variability related to blood pressure regulation include oscillatory catecholamine levels, vascular adrenergic reactivity, baro-reflex function, ^23, 24^ renin-angiotensin aldosterone system activity, ^25^ cardiac metabolism and response to pro-hypertrophic stimuli.^26, 27^

A growing body of work suggests that dampening or loss of amplitude in oscillatory processes are associated with aging,^28^ age-dependent diseases,^29, 30^ and elevated disease risk.^31^ Our findings are in agreement with this framework; however, future efforts are necessary for validation where mechanistic insight drives clinical application. Mouse models with genetically engineered dysfunctional molecular clocks exhibited altered 24-hour BP profiles.^32^ Vascular stiffening and impaired vascular remodeling have been implicated with a dysregulated matrix metalloproteinase pathway in animal models of clock disruption.^33^ This mechanism of accelerated hypertension-mediated end organ damage,^34^ is a likely factor contributing to the increased risk of death from cardiovascular causes that we detected in the CRIC participants without cyclic blood pressure components.

Wearable cuffless blood pressure devices are being developed and some are currently on the market. ^35^ These devices aim to provide repeat assessments over longer periods of time, are less disruptive to patients’ sleep and daily activities, and may provide a better reproducible alternative to current cuff-based ABPMs. Refined measures of diurnal blood pressure variation as presented in this study are a powerful tool to identify patients at excess risk of disease.

Our study has several strengths. We used two well designed cohorts that characterized participants extensively and followed them up for sufficient periods of time. Both studies also applied standardized procedures for both ABPM measurements and for the adjudication of outcomes. While the sample size is large in the CRIC cohort, the number of participants and events remains smaller in the AASK cohort which limits the power to detect a signal above the noise. Compared to CRIC, the event rate in AASK for cardiovascular death was much lower, suggesting that the cohort disease structure is substantially different. This may be the reason why we were not able to reproduce our CRIC findings among AASK participants. The value, however, lies in synthesizing both approaches to define future clinical study designs. A crucial difference is the high proportion of diabetics in CRIC (42%) compared to AASK (13%). Circadian autonomic dysfunction, a frequent observation among diabetics,^36^ could be responsible for an increase in effect size with subsequent detection of a signal above the noise in CRIC but not in AASK. This information could rationalize an initial focus on CKD patients with concomitant cardiovascular and diabetic comorbidities. Also, at the trial phase, the AASK study excluded non-hypertensive kidney disease patients and diabetics. Many of the CRIC cohort patients would not have qualified for participation in the AASK making these two cohorts unique. Furthermore, the high prevalence of CVD among participants who died due to cardiovascular causes (60 out of 83) necessitated a subgroup analysis in patients with prior cardiovascular disease to investigate the association between the loss of rhythmic components and cardiovascular death beyond that which is confounded by prior cardiovascular disease. This decreased the number of cardiovascular deaths in this subgroup analysis leading to a wide confidence interval for our significant association with cardiovascular death. Our dipping category subgroup analysis is also limited by the decreased resulting event numbers, however it provides necessary results that further differentiate rhythmic components from a calculation of the nocturnal dip. Sleep-disordered breathing such as sleep apnea is known to disrupt sleep-wake rhythms and should also be considered as a source to dampen rhythmic blood pressure components. We were not able to examine this in the present study.

### Non-Standard Abbreviations

ABPM: ambulatory blood pressure monitoring
ARV: average real variability
BMAL1: brain and muscle ARNT-like protein 1
DR: dipping ratio
MAP: mean arterial pressure

## Data Availability

All data produced in the present work are contained in the manuscript

## Acknowledgments

We thank the NIDDK-CR and the CRIC Publication Executive Committee for making the data available for our use in this study. CRIC Study Investigators are in alphabetical order Lawrence J. Appel, MD, MPH, Jing Chen, MD, MMSc, MSc, Debbie L Cohen, MD, Harold I. Feldman, MD, MSCE, Alan S. Go, MD, James P. Lash, MD, Mahboob Rahman, MD, and Mark L. Unruh, MD, MS.

## Supplementary Material

Figures S1-S3

Tables S1-S7

## Supplementary Figures

**Figure S1.**
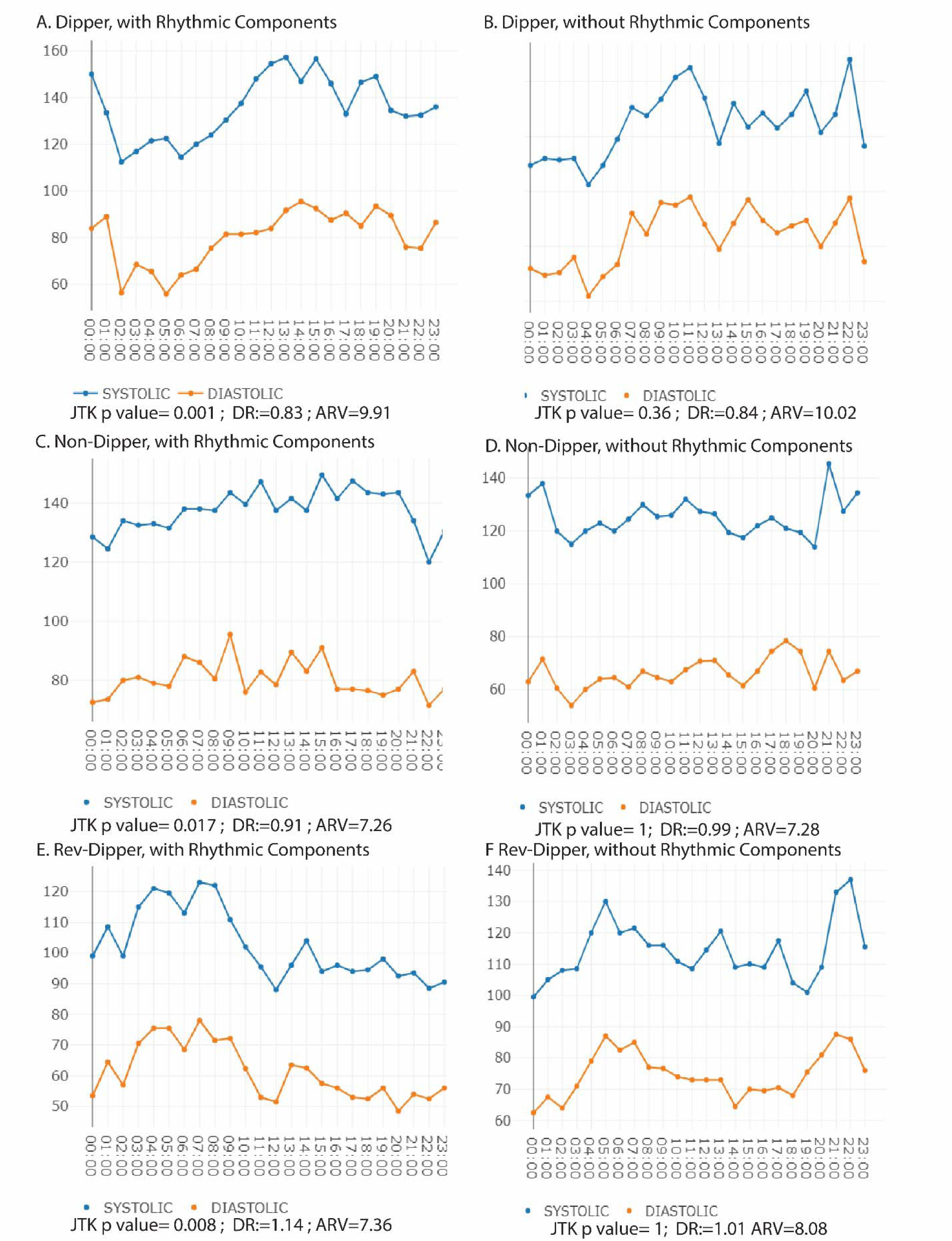
Juxtaposition of 24 hour blood pressure curves and blood pressure variability metrics from six CRIC participants to illustrate the discriminatory characteristics on the basis of the JTK_CYCLE analysis while **DR (**Dipping Ratio) and **ARV (**Average real variability) show similar values between pairs of dippers in A and B, non-dippers in C and D, and reverse dippers in E and F.

**Figure S2.**
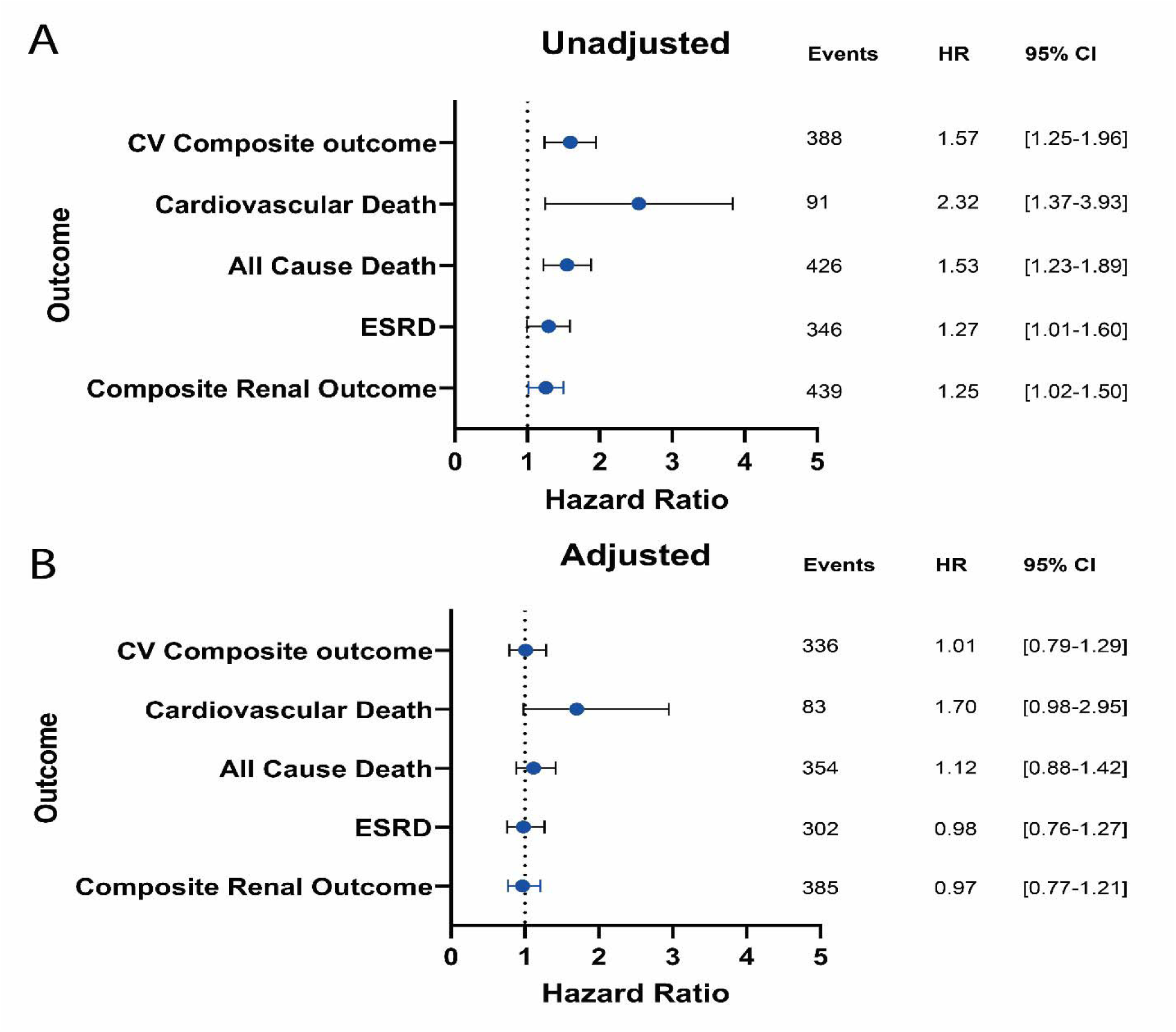
Hazard ratios for the absence of rhythmic components (JTK p value >0.05) and reaching different outcomes in the CRIC cohort as compared to the retention of rhythmic components (JTK p value ≤0.05). **A.** Unadjusted model **B.** Adjusted for Age, BMI, Sex, Diabetes, Race, eGFR, Urine Protein to Creatinine Ratio, Clinic SBP, Clinic DBP, Prior CVD.

**Figure S3:**
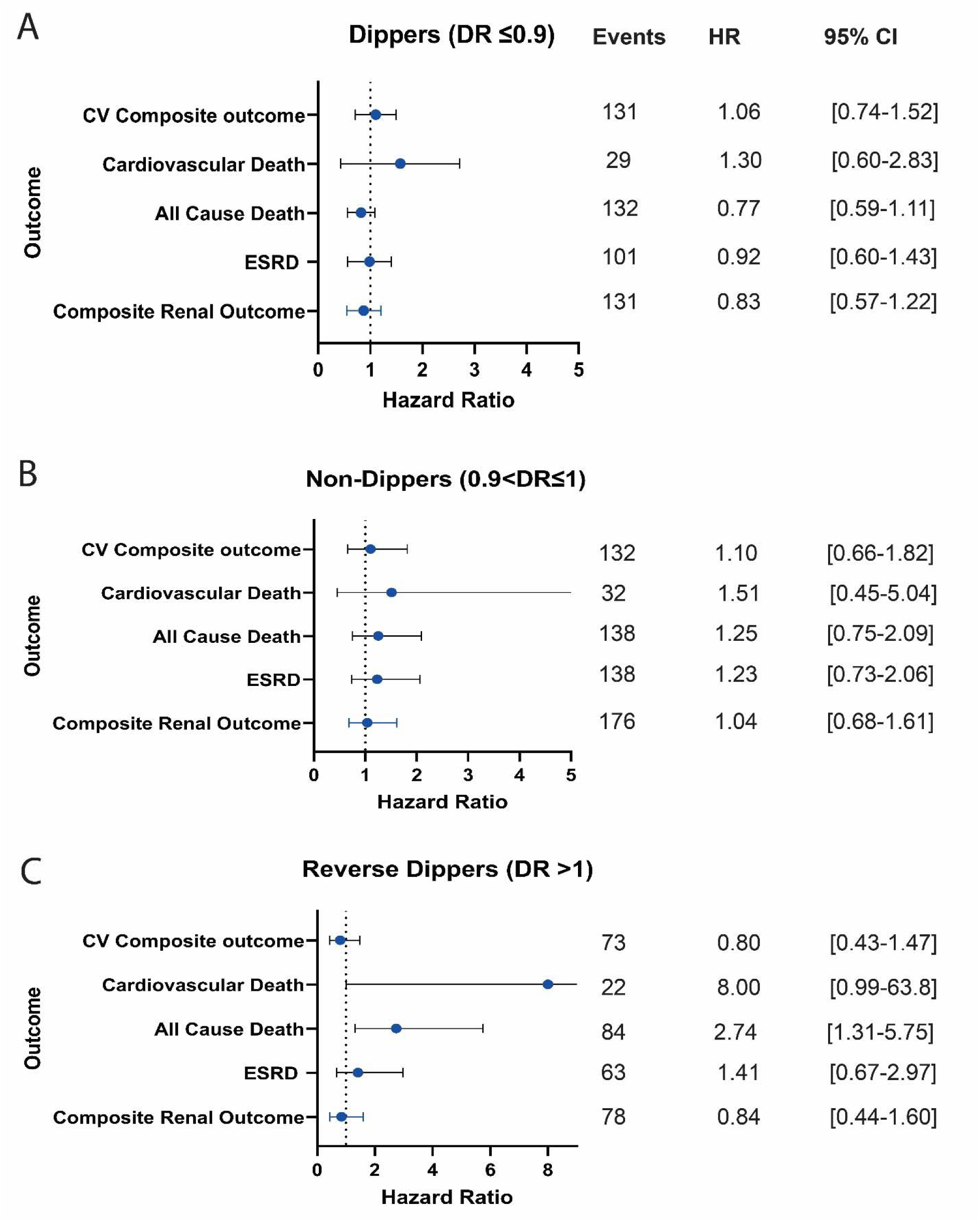
Hazard ratios for the absence of rhythmic components (JTK p value >0.05) and reaching different outcomes as compared to the retention of rhythmic components (JTK p value ≤0.05) from the CRIC cohort. Adjusted for Age, BMI, Sex, Diabetes, Race, eGFR, Urine Protein to Creatinine Ratio, Clinic SBP, Clinic DBP, Prior CVD. **A.** Analysis performed only in dippers. **B.** Analysis performed only in non-dippers. **C.** Analysis performed only in reverse dippers. DR: Dipping ratio, CV: Cardiovascular, CVD: Cardiovascular disease, ESRD: End stage renal disease.

## Supplemental Tables

**Table S1.** Blood pressure profiles of participants in both cohorts. Blood pressure parameters were computed from 24 Hour ABPM (the first 24 Hour ABPM in AASK).

|  | **CRIC** | **AASK** |
| --- | --- | --- |
|  | **N(%)** | **N(%)** |
| **Rhythmic Components present (JTK $p \leq 0.05$)** | 514 (34%) | 169 (26%) |
| **Rhythmic Components absent (JTK $p > 0.05$)** | 988 (66%) | 474 (74%) |
| **Dipper (DR $\leq 0.9$)** | 678 (45%) | 127 (20%) |
| **Dipper w/ rhythmic components (DR $\leq 0.9$ & JTK $< 0.05$)** | 376 (25%) | 54 (8%) |
| **Non-Dipper (0.9 $<$ DR $\leq 1$)** | 572 (38%) | 266 (41%) |
| **Non-Dipper w/ rhythmic components (0.9 $<$ DR $\leq 1$ & JTK $< 0.05$)** | 92 (6%) | 38 (6%) |
| **Reverse Dipper (DR $> 1$)** | 252 (17%) | 250 (39%) |
| **Reverse Dipper w/ rhythmic components (DR $> 1$ & JTK $< 0.05$)** | 46 (3%) | 77 (12%) |
| **Controlled BP (24 Hr Mean $< 125/75$)** | 549 (37%) | 100 (16%) |
| **Uncontrolled BP (24 Hr Mean ≥125/75)** | 953 (63%) | 543 (85%) |

**Table S2.** Multivariate logistic regression for non-dipping status (dipping ratio >0.9) among CRIC cohort participants. All variables in the table were included as covariates in the model. eGFR: estimated glomerular filtration rate by the CRIC cohort equation. BP: blood pressure. PCR: urine protein to creatinine ratio.

| **Covariate** | **Odds**<br>**Ratio** | **95%CI**<br>**Lower** | **95%CI**<br>**Upper** | **P-value** |
| --- | --- | --- | --- | --- |
| Age <45 | Reference |  |  |  |
| Age From 45 to <65 | 1.32 | 0.8 | 2.18 | 0.276 |
| Age 65+ | 1.17 | 1.01 | 2.78 | 0.047 |
| BMI <25 | Reference |  |  |  |
| BMI 25 to <30 | 1.07 | 0.74 | 1.55 | 0.705 |
| BMI ≥30 | 1.27 | 0.89 | 1.8 | 0.182 |
| eGFR >60 ml/min/1.73m<sup>2</sup> | Reference |  |  |  |
| eGFR: 30 to <60 ml/min/1.73m<sup>2</sup> | 1.11 | 0.82 | 1.51 | 0.483 |
| eGFR<30 ml/min/1.73m<sup>2</sup> | 1.46 | 0.99 | 2.15 | 0.059 |
| Proteinuria (PCR <150mg/g) | Reference |  |  |  |
| Proteinuria (PCR: 150-500 mg/g) | 1.08 | 0.8 | 1.46 | 0.623 |
| Proteinuria (PCR>500 mg/g) | 1.56 | 1.12 | 2.17 | 0.009 |
| Controlled BP | Reference |  |  |  |
| Un-Controlled BP (Mean 24 Hr<br>BP ≥ 125/75mmHg) | 1.64 | 1.27 | 2.12 | <0.001 |
| Male Sex | Reference |  |  |  |
| Female Sex | 0.85 | 0.67 | 1.09 | 0.195 |
| Non-Diabetic | Reference |  |  |  |
| Diabetic | 1.23 | 0.96 | 1.67 | 0.1 |
| Race: White | Reference |  |  |  |
| Race: Black | 1.55 | 1.2 | 2.01 | 0.001 |
| Race: Other | 0.92 | 0.63 | 1.32 | 0.64 |
| Prior CVD: No | Reference |  |  |  |
| Prior CVD: Yes | 1.58 | 1.23 | 2.03 | <0.001 |

**Table S3.**
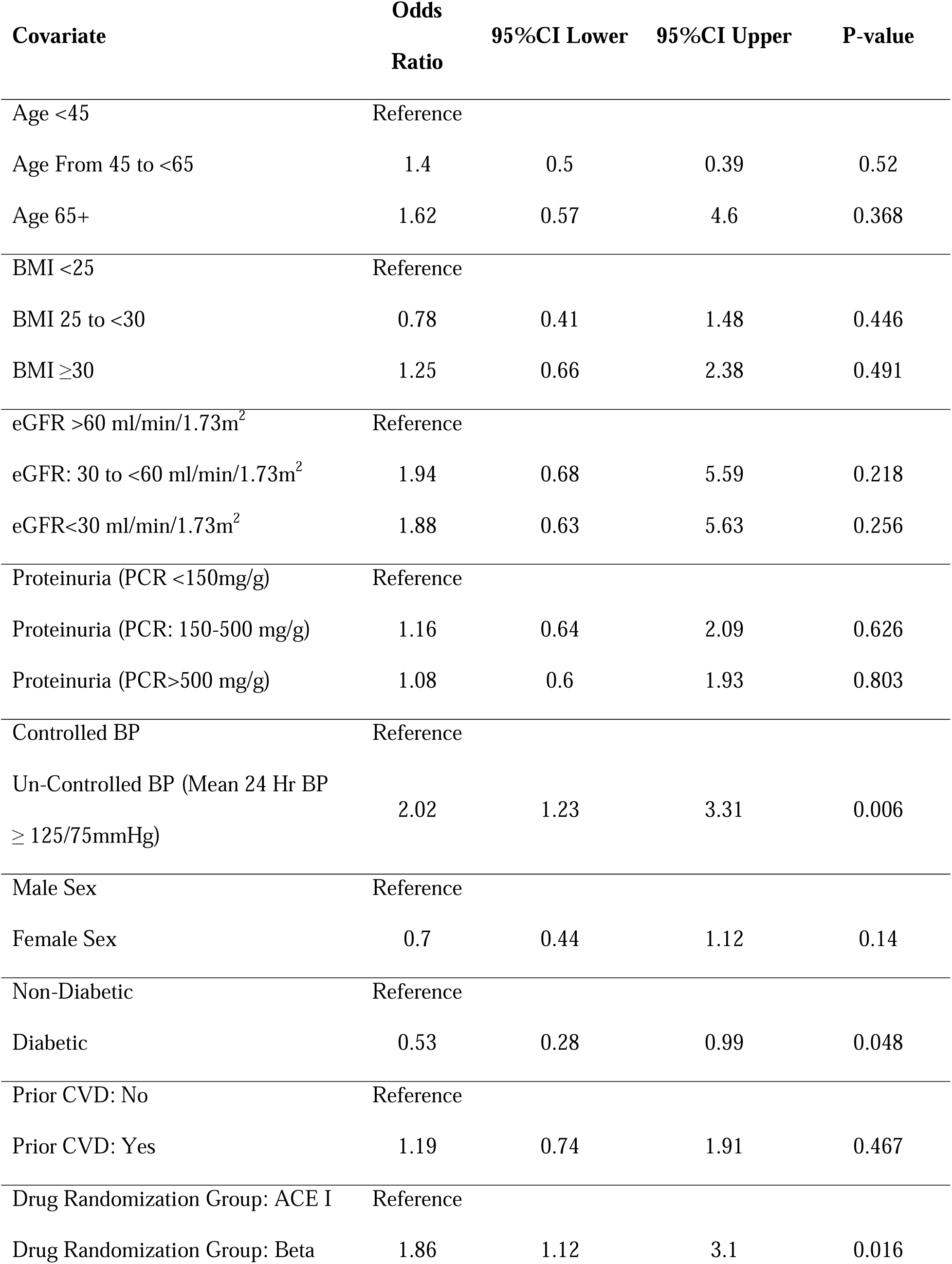
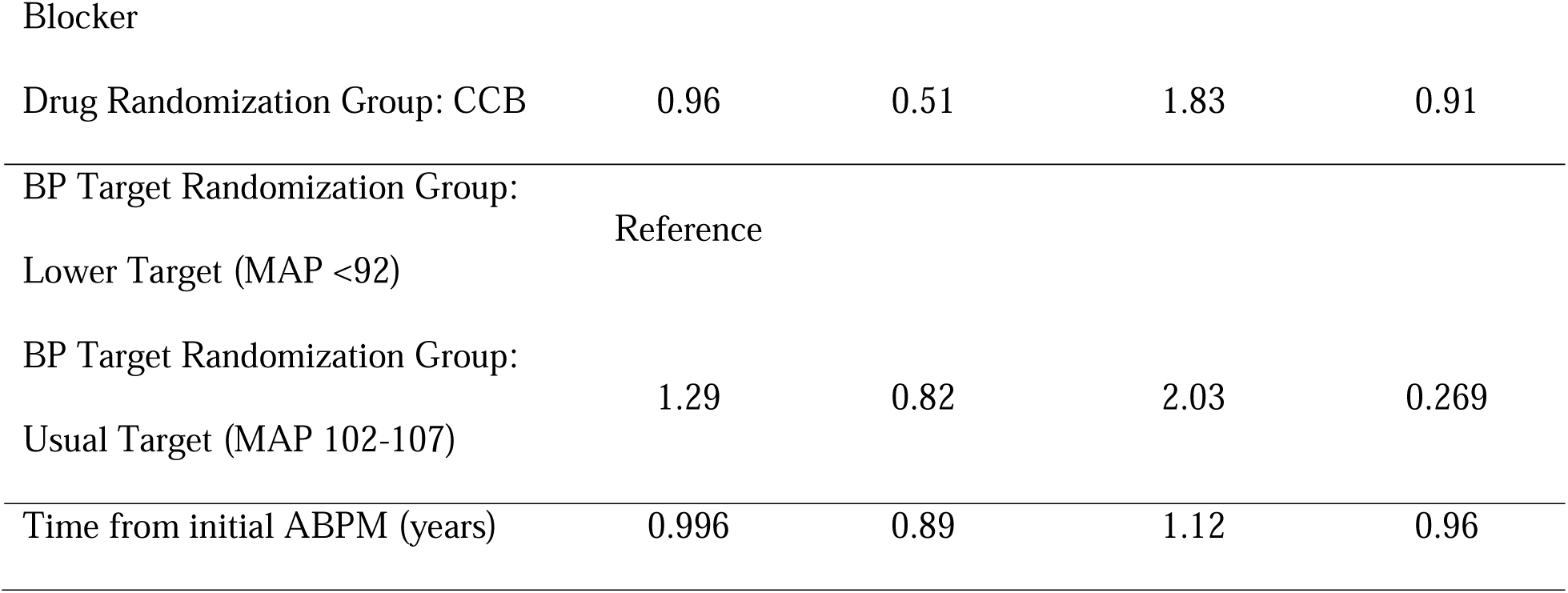
Mixed effects logistic regression for non-dipping status (dipping ratio >0.9 among AASK cohort participants. All variables in the table were included as covariates in the model. eGFR: estimated glomerular filtration rate by the 2021 CKD-EPI equation. PCR: urine protein to creatinine ratio. BP: blood pressure. ACE I: angiotensin converting enzyme inhibitor. CCB: calcium channel blocker.

**Table S4.**
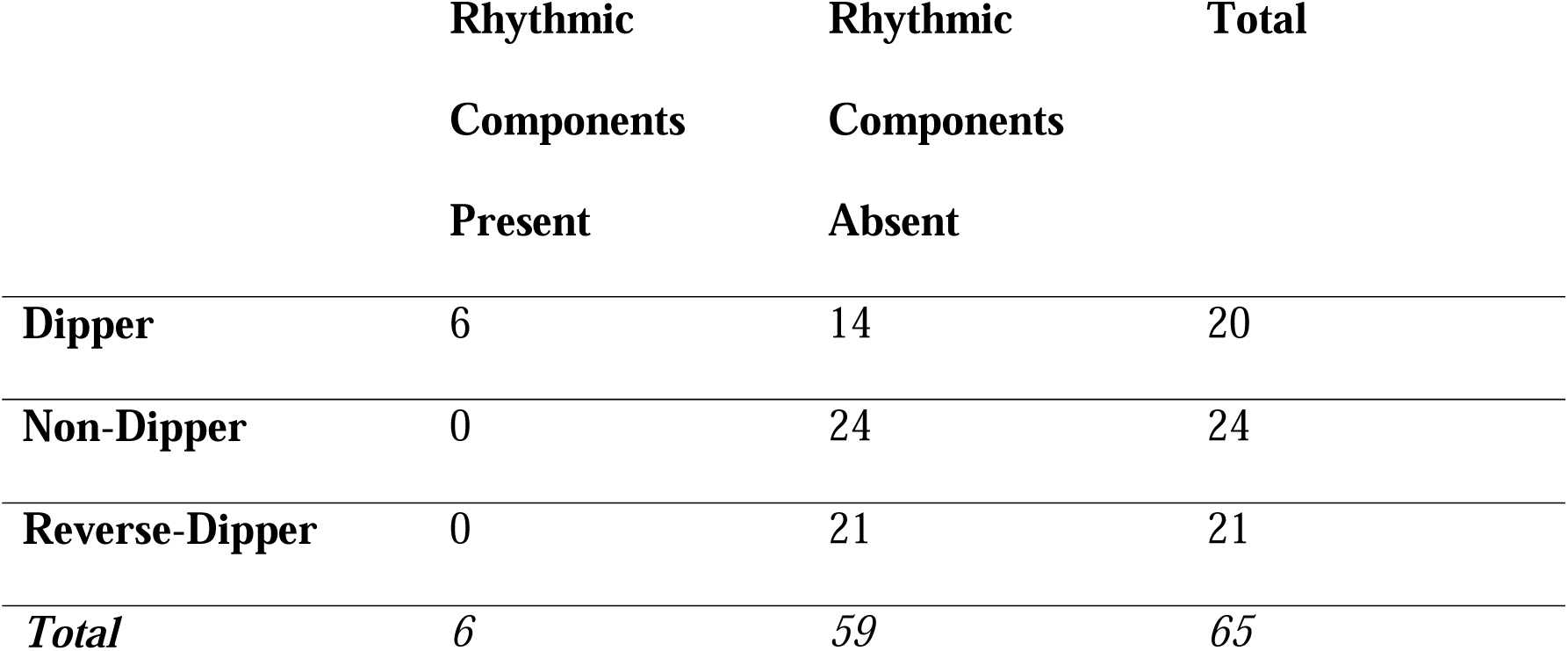
Cross Tabulation of dipping status and the presence or absence of rhythmic components in ABPM among participants with prior cardiovascular disease who died due to cardiovascular causes.

|  | **Rhythmic**<br>**Components**<br>**Present** | **Rhythmic**<br>**Components**<br>**Absent** | **Total** |
| --- | --- | --- | --- |
| **Dipper** | 6 | 14 | 20 |
| **Non-Dipper** | 0 | 24 | 24 |
| **Reverse-Dipper** | 0 | 21 | 21 |
| ***Total*** | **6** | **59** | **65** |

**Table S5.** Hazard ratios for the different dipping categories and reaching different outcomes from the CRIC cohort. Adjusted for Age, BMI, Sex, Diabetes, Race, eGFR, Urine Protein to Creatinine Ratio, Clinic SBP, Clinic DBP, Prior CVD. DR: Dipping ratio, CV: Cardiovascular, CVD: Cardiovascular disease, ESRD: End stage renal disease.

| Outcome | Dipping Category | Unadjusted Model |  |  | Adjusted Model |  |  |
| --- | --- | --- | --- | --- | --- | --- | --- |
|  |  | Hazard Ratio | 95% CI | p-value | Hazard Ratio | 95% CI | p-value |
| **Composite Renal Outcome** | Dipper | Reference |  |  |  |  |  |
|  | Non-Dipper | 1.81 | 1.47-2.23 | <0.001 | 1.27 | 1.01-1.61 | 0.04 |
|  | Reverse Dipper | 2.99 | 1.53-2.60 | <0.001 | 1.40 | 1.05-1.90 | 0.021 |
| **ESRD** | Dipper | Reference |  |  |  |  |  |
|  | Non-Dipper | 1.8 | 1.42-2.28 | <0.001 | 1.07 | 0.82-1.41 | 0.617 |
|  | Reverse Dipper | 1.97 | 1.47-2.66 | <0.001 | 1.37 | 0.98-1.91 | 0.064 |
| **All Cause Death** | Dipper | Reference |  |  |  |  |  |
|  | Non-Dipper | 1.4 | 1.12-1.74 | 0.003 | 0.93 | 0.73-1.18 | 0.549 |
|  | Reverse Dipper | 2.44 | 1.91-3.12 | <0.001 | 1.14 | 0.85-1.53 | 0.373 |
| **Cardiovascular Death** | Dipper | Reference |  |  |  |  |  |
|  | Non-Dipper | 1.3 | 0.80-2.12 | 0.297 | 0.98 | 0.59-1.63 | 0.931 |
|  | Reverse Dipper | 2.55 | 1.52-4.28 | <0.001 | 1.12 | 0.61-2.04 | 0.717 |

|  | Dipper | Reference |  |  |  |  |  |
| --- | --- | --- | --- | --- | --- | --- | --- |
| **CV Composite** | Non-Dipper | 1.35 | 1.07-1.69 | 0.01 | 0.92 | 0.72-1.18 | 0.501 |
| **outcome** | Reverse Dipper | 2.09 | 1.61-2.72 | <0.001 | 1.01 | 0.75-1.36 | 0.951 |

**Table S6.**
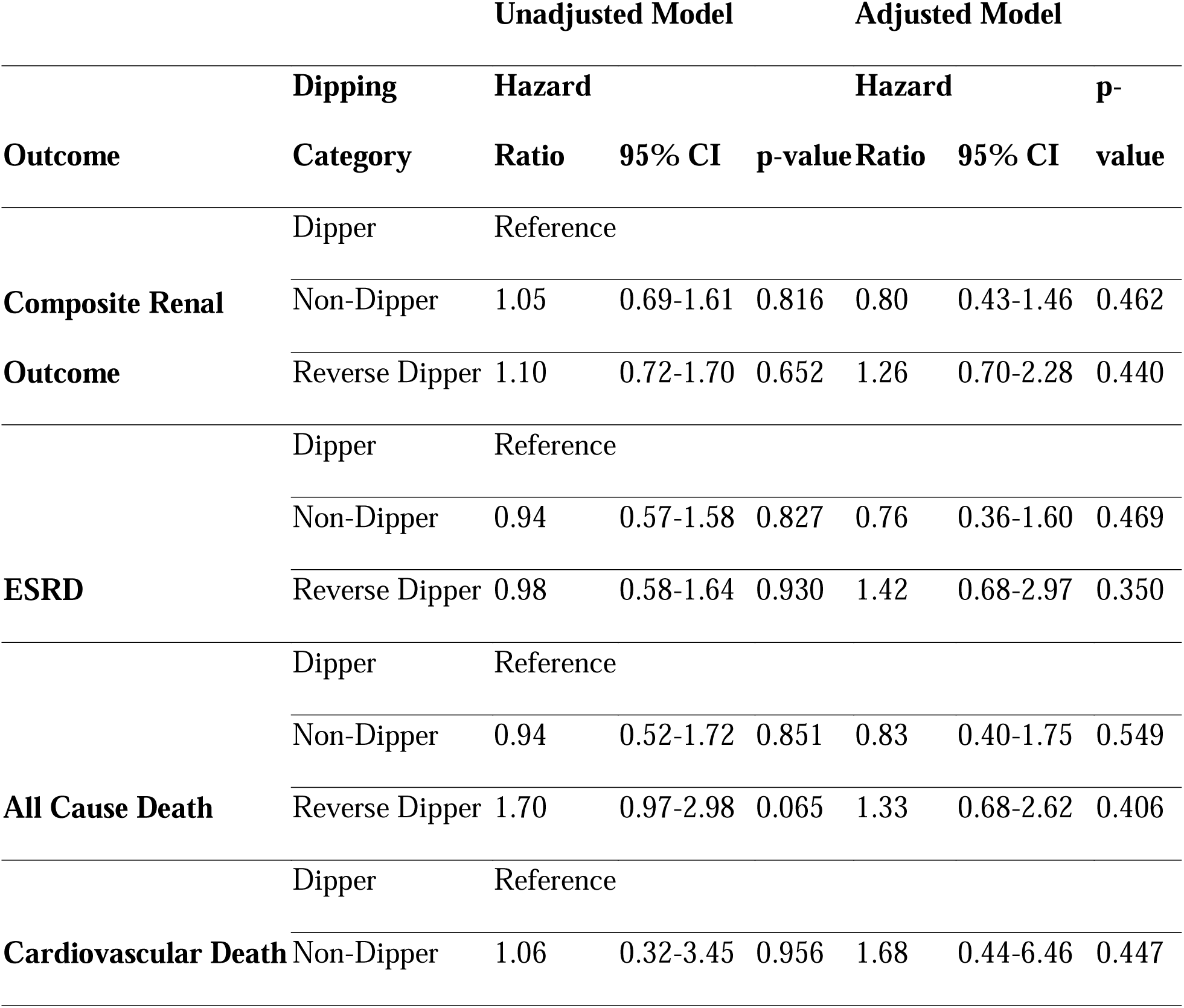
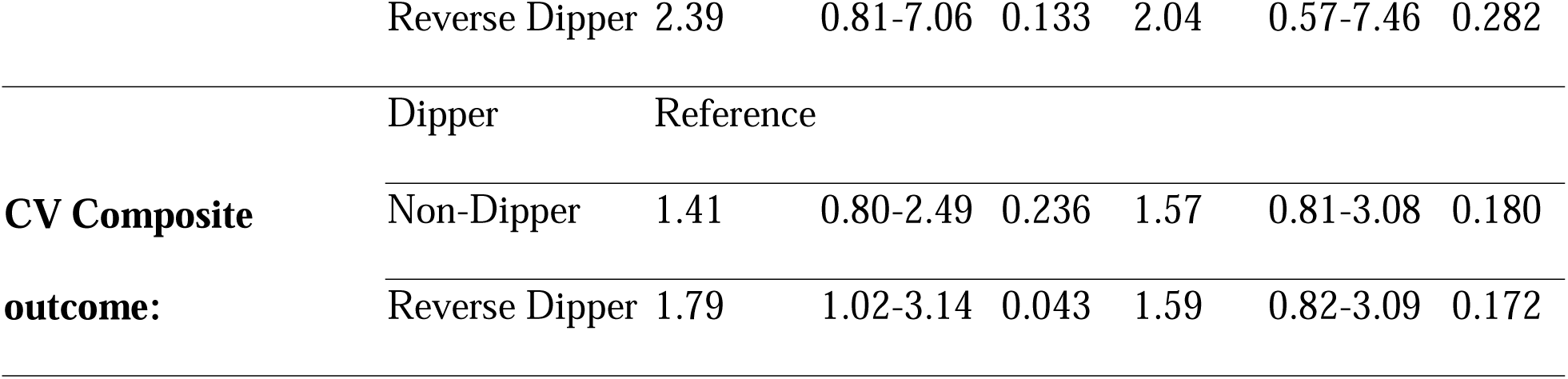
Hazard ratios for the different dipping categories and reaching different outcomes from the AASK cohort. Adjusted for Age, BMI, Sex, Diabetes, eGFR, Urine Protein to Creatinine Ratio, Clinic SBP, Clinic DBP, Prior CVD Drug and blood pressure target groups randomized to in the prior trial. DR: Dipping ratio, CV: Cardiovascular, CVD: Cardiovascular disease, ESRD: End stage renal disease.

**Table S7.**
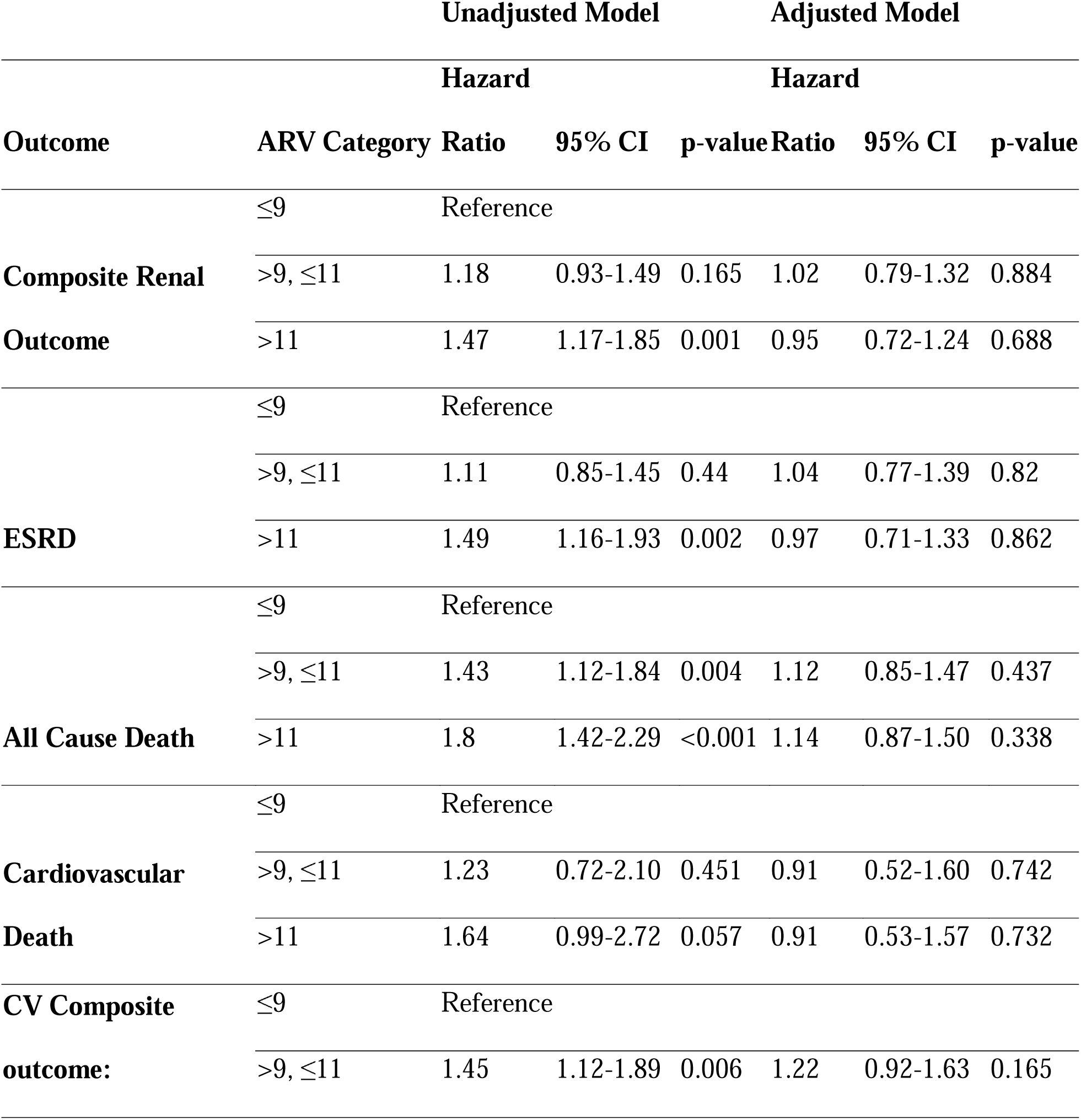
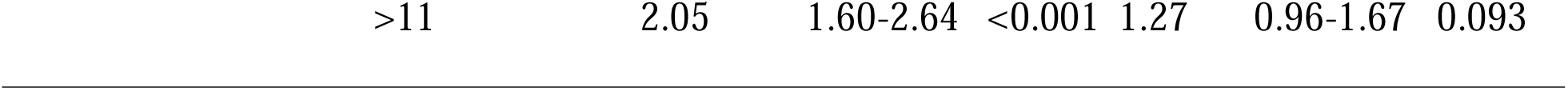
Hazard ratios for the different average real variability (ARV) tertials and reaching different outcomes from the CRIC cohort. Adjusted for Age, BMI, Sex, Diabetes, Race, eGFR, Urine Protein to Creatinine Ratio, Clinic SBP, Clinic DBP, Prior CVD. CV: Cardiovascular, CVD: Cardiovascular disease, ESRD: End stage renal disease.

